# Post COVID-19 Condition in South Africa: 3-month follow-up after hospitalisation with SARS-CoV-2

**DOI:** 10.1101/2022.03.06.22270594

**Authors:** Murray Dryden, Caroline Mudara, Caroline Vika, Lucille Blumberg, Natalie Mayet, Cheryl Cohen, Stefano Tempia, Arifa Parker, Jeremy Nel, Rubeshan Perumal, Michelle J. Groome, Francesca Conradie, Norbert Ndjeka, Louise Sigfrid, Laura Merson, Waasila Jassat

**Author notes:** **Corresponding author:** Dr Murray Dryden, Division of Public Health Surveillance and Response, National Institute for Communicable Diseases, 1 Modderfontein Road Sandringham.

## Abstract

**Background:** Post COVID-19 Condition (PCC) as defined by WHO refers to a wide range of new, returning, or ongoing health problems experienced by COVID-19 survivors, and represents a rapidly emerging public health priority. We aimed to establish how this developing condition has impacted patients in South Africa and which population groups are at risk.

**Methods:** In this prospective cohort study, participants ≥18 years who had been hospitalised with laboratory-confirmed SARS-CoV-2 infection during the second and third wave between December 2020 and August 2021 underwent telephonic follow-up assessment up at one-month and three-months after hospital discharge. Participants were assessed using a standardised questionnaire for the evaluation of symptoms, functional status, health-related quality of life and occupational status. Multivariable logistic regression models were used to determine factors associated with PCC.

**Findings:** In total, 1,873 of 2,413 (78%) enrolled hospitalised COVID-19 participants were followed up at three-months after hospital discharge. Participants had a median age of 52 years (IQR 41-62) and 960 (51.3%) were women. At three-months follow-up, 1,249 (66.7%) participants reported one or more persistent COVID-related symptom(s), compared to 1,978/2,413 (82.1%) at one-month post-hospital discharge. The most common symptoms reported were fatigue (50.3%), shortness of breath (23.4%), confusion or lack of concentration (17.5%), headaches (13.8%) and problems seeing/blurred vision (10.1%). On multivariable analysis, factors associated with new or persistent symptoms following acute COVID-19 were age ≥65 years [adjusted odds ratio (aOR) 1.62; 95%confidence interval (CI) 1.00-2.61]; female sex (aOR 2.00; 95% CI 1.51-2.65); mixed ethnicity (aOR 2.15; 95% CI 1.26-3.66) compared to black ethnicity; requiring supplemental oxygen during admission (aOR 1.44; 95% CI 1.06-1.97); ICU admission (aOR 1.87; 95% CI 1.36-2.57); pre-existing obesity (aOR 1.44; 95% CI 1.09-1.91); and the presence of ≥4 acute symptoms (aOR 1.94; 95% CI 1.19-3.15) compared to no symptoms at onset.

**Interpretation:** The majority of COVID-19 survivors in this cohort of previously hospitalised participants reported persistent symptoms at three-months from hospital discharge, as well as a significant impact of PCC on their functional and occupational status. The large burden of PCC symptoms identified in this study emphasises the need for a national health strategy. This should include the development of clinical guidelines and training of health care workers, in identifying, assessing and caring for patients affected by PCC, establishment of multidisciplinary national health services, and provision of information and support to people who suffer from PCC.

## Introduction

While there is a growing understanding of acute coronavirus disease 2019 (COVID-19) and risk factors for severe disease and death, less is known about ongoing and long-term complications. There has been debate about the most appropriate nomenclature and clinical criteria for the long-term complications of COVID-19, with various terminology used over time, including Long COVID, Long Haulers, and Post-Acute Sequelae of COVID-19 (PASC). The World Health Organization (WHO) conducted a global Delphi study to arrive at a consensus on the name and clinical definition of the condition. Post COVID-19 Condition (PCC) “occurs in individuals with a history of probable or confirmed SARS-CoV-2 infection, usually 3 months from the onset of COVID-19 with symptoms that last for at least 2 months and cannot be explained by an alternative diagnosis” (1). WHO also clarifies that the symptoms may be new onset, following initial recovery from an acute COVID-19 episode, or persist from the initial illness and that they may fluctuate or relapse over time.

Cases of persistent symptoms following acute COVID-19 were described as early as May 2020, and our understanding of the condition continues to improve through research. Initial estimates suggested that 1 in 10 people (2) infected with SARS-CoV-2 will experience new or persistent symptoms beyond four weeks of their acute illness, irrespective of the initial disease severity. A recent systematic review by Groff *et al*. (3), which included 57 studies, reported pulmonary sequelae, neurologic disorders, mental health disorders, functional mobility impairments, and general and constitutional symptoms. About half of patients experienced PASC for more than six-months (4). The most prevalent sequelae included impaired concentration, generalised anxiety disorder, fatigue, and muscle weakness and functional mobility disorder. Another study found that at seven months post-COVID-19 onset, 45% had not returned to previous levels of work, and continued to experience significant symptom burden (5).

The frequency, clinical picture and impact of PCC could vary in different settings as a result of different patient genetic and social background. The impact of PCC could be worse in Africa because people often have unreliable income and poor access to health services. Serology studies suggest that in South Africa >70% of people have experienced SARS-CoV-2 infection (6) but to our knowledge there are no published studies to describe PCC. As part of a multi-country study coordinated by the International Severe Acute Respiratory and emerging Infection Consortium (ISARIC), we established a prospective cohort of SARS-CoV-2 infected participants for serial follow-up after hospitalisation to determine the prevalence of and risk factors for PCC among individuals hospitalised with laboratory-confirmed SARS-CoV-2 infection in South Africa.

## Method

### Study design

This was a prospective, observational cohort study using an ISARIC open-access tool that was locally adapted to follow-up participants with COVID-19 in South Africa (7).

### Study population

The DATCOV national hospital surveillance system developed by the National Institute for Communicable Diseases (NICD) and the National Department of Health (NDoH), was used to identify SARS-CoV-2-infected hospitalised participants and to obtain baseline patient data on demographic characteristics, comorbidities and hospital admissions. The study population included hospitalised individuals in the public and private health sectors from all provinces of South Africa. Patients with a positive reverse transcriptase polymerase chain reaction (RT-PCR) assay or rapid antigen test for SARS-CoV-2, hospitalised for a minimum of one day, and discharged between the period 1 December 2020 to 27 July 2021. We did not routinely perform viral sequencing to establish infecting SARS-CoV-2 variants, however based on South African sequencing data the two most common variants during the study period were the Beta (epidemiologic week 1 to 20) and Delta variants (epidemiologic week 21 to 30) (8). Ethnicity was determined by self-identification and categorised in line with the Statistics South Africa classification, including black, white, mixed, Indian, and other.

Participants were eligible for inclusion irrespective of reason for admission, including those admitted due to COVID-19 or with SARS-CoV-2 infection identified during the hospitalisation. Of all eligible, discharged, adult participants with contact details available, a random selection of participants were invited by telephone for participation in this study. Random sampling was performed using a computer-generated list of eligible participants. Participants 18 years and older who consented verbally to participate in the study were included.

### Measurements instruments

A standardised case report form (CRF) and follow-up protocol developed by ISARIC in collaboration with members of the global Long COVID support group was used for demographic variables, COVID-19 vaccination status, symptoms during hospitalisation, current health status, acute COVID-19 complications, new or persistent symptoms, lifestyle and socioeconomic variables. The CRF was piloted with patients in three different countries before it was finalised. The CRF contained validated tools to establish quality of life (measured by EQ-5D-5L) (9), dyspnoea (assessed using modified MRC dyspnoea scale) (10) and difficulties in functioning (UN/Washington disability score) (11). Changes in disability, breathlessness and health state (EQ5D index) were calculated by comparing participants before SARS-CoV-2 infection to the three-month follow-up assessment. The Washington Group Short Set on Functioning (WG-SS) questions (11) were used to measure changes in short term disability (seeing, hearing, walking, remembering, communication and self-care).

### Data collection

Participants were initially enrolled by means of a telephonic assessment at one-month after hospital discharge, with additional scheduled follow-up assessments at three-, six- and 12-months post-hospital discharge. Verbal consent was obtained and recorded, and, where possible, interviews were conducted in the language of the participants’ choice (English, isiZulu, isiXhosa, SeSotho and Afrikaans). Data was entered and stored on a secure online Research Electronic Data Capture (REDCap, version 10.6.14, Vanderbilt University, Nashville, Tenn.) repository hosted by the University of Oxford on behalf of ISARIC.

### Statistical analysis

This paper reports the prevalence of symptoms at one- and three-month follow-up and presents a detailed analysis of PCC at three months from hospital discharge.

Frequencies and percentages were used to summarise categorical data, and continuous data were expressed using medians and interquartile ranges (IQR). Frequency distribution tables and graphs were used to describe demographics, the prevalence of symptoms, comorbidities, and changes in health and occupational status.

Five multivariable logistic regression models were implemented to assess factors associated with the following outcomes at three months from hospital discharge: 1) new or persistent symptoms, 2) self-reported non-recovery, 3) new or worsening breathlessness, 4) new or worsening disability, and 5) depression/anxiety (Supplementary Methods for description of how outcome variables were defined).

Predictor variables assessed for association with the different outcomes in the multivariable models were age, sex, ethnicity, presence of individual comorbid conditions (asthma, diabetes, hypertension, chronic cardiac disease, chronic kidney disease, malignancy, tuberculosis, HIV and obesity), number of symptoms during acute infection, treatment in ICU and treatment with oxygen or ventilation. For each multivariable model, variables with p<0.2 in the univariate analysis were included for multivariate analysis. Some variables such as age, sex and ethnicity were included in the model *a priori* based on clinical plausibility. Manual backward elimination was implemented and the final model selection was guided by minimisation of the Akaike information criterion (AIC) or Bayesian information criterion (BIC). Statistical significance for the multivariable analysis was assessed at p<0.05. Statistical analyses were performed using Stata software version 16 (StataCorp Limited, College Station, Texas, USA).

### Ethics and approvals

The study was approved by the University of the Witwatersrand Human Research Ethics Committee (HREC M201150). Approvals were obtained from all provinces via the National Health Research Database (NHRD).

## Results

### Study population

Of the 241,159 COVID-19 admissions reported to DATCOV between 1 December 2020 and 23 August 2021, 129,591 were eligible for inclusion and 8,312 were randomly selected for enrolment. Of the 3,097 patients that we managed to reach, 2,413 (77.9%) consented to participate in the study (Figure 1). The enrolled participants and those who were not enrolled in the study had similar demographic characteristics and distribution of comorbidities, except that they differed by the health sector they were hospitalised in (more patients admitted in the private sector included) (Supplementary Table 1).

**Figure 1.**
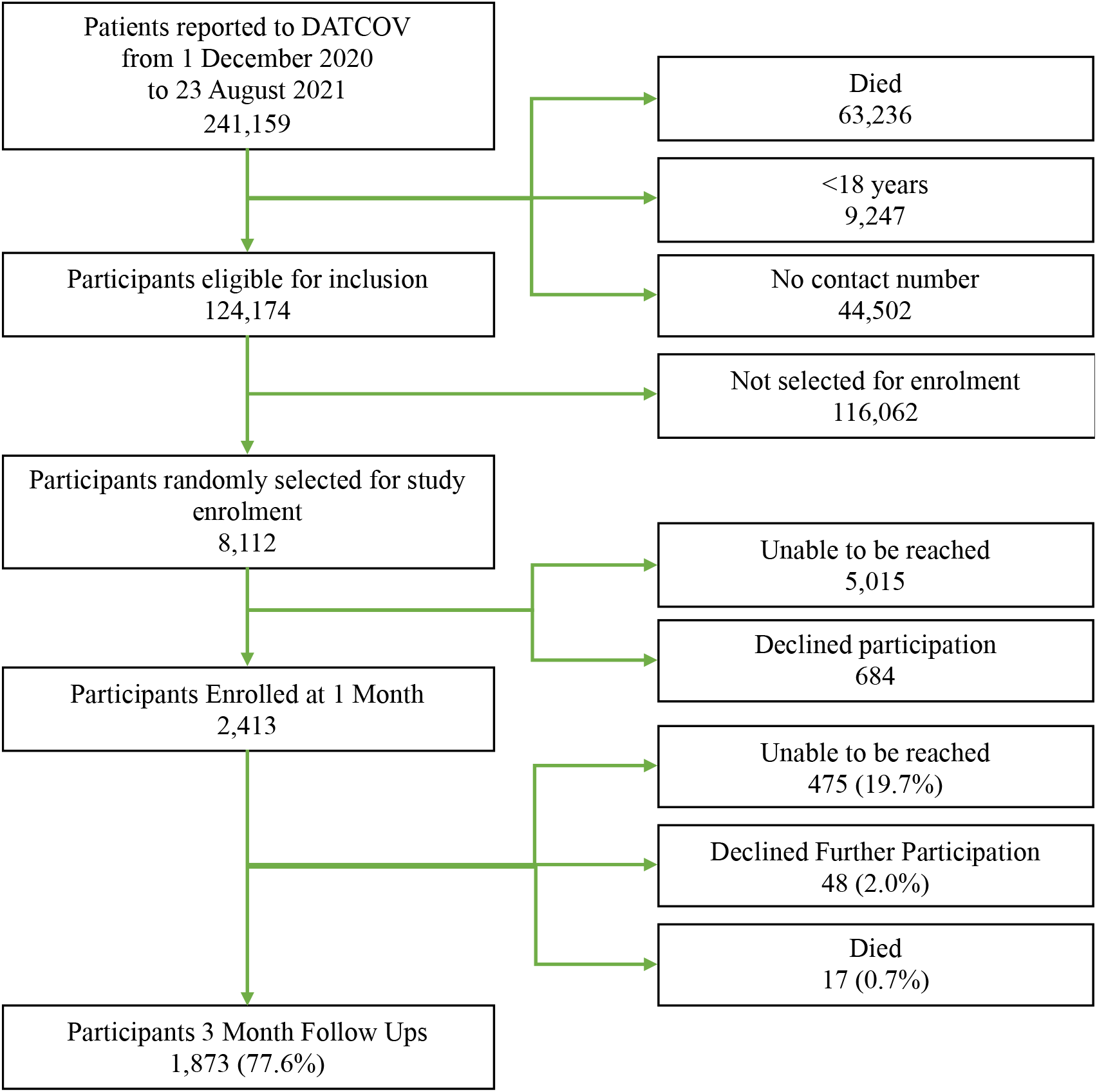
Study population based on inclusion criteria, PCC Study, South Africa

Among the participants followed up at three months, the median time between hospital discharge and telephonic follow-up was 95 days (IQR 84-126). Of the 1,873 participants, 960 (53.1%) were female. The median age was 52 years (IQR 41-62) and most participants were aged 40-64 years (n=1,102; 58.8%). There were 896 (47.8%) participants who were black, 677 (36.2%) white, 154 (8.2%) mixed and 132 (7.1%) Indian (Table 1). More than two thirds of the participants had at least one comorbid condition (n=1,316; 70.3%). The most common self-reported comorbidities were hypertension (n=669; 35.7%), obesity (n=474; 25.3%) and diabetes mellitus (n=418; 22.3%). HIV was reported by 95 (5.1%) participants. There were 462 (24.7%) participants hospitalised in the public sector. The majority (n=1,324; 70.7%) of participants received supplemental oxygen therapy during hospitalisation, whilst 612 (32.7%) required admission to an intensive care unit (ICU) and 179 (9.6%) received invasive mechanical ventilation (Table 1).

**Table 1.** Characteristics of participants at 3 months follow-up in the PCC study, South Africa

| Characteristics | Female | Male | Total |
| --- | --- | --- | --- |
|  | 960 (51.3%) | 913 (48.8%) | N=1873 |
| **Median age (IQR)** | 50 [38 - 60] | 53 [44 -63] | 52 [41 - 62] |
| **Age group** |  |  |  |
| <40 years | 226 (16.0) | 181 (39.2) | 407 (21.7) |
| 40-64 years | 880 (62.4) | 222 (48.1) | 1,102 (58.8) |
| ≥65 years | 305 (21.6) | 59 (12.8) | 364 (19.4) |
| **Ethnicity** |  |  |  |
| Black | 515 (53.7) | 381 (41.7) | 896 (47.8) |
| White | 306 (31.9) | 371 (40.6) | 677 (36.2) |
| Mixed | 73 (7.6) | 82 (8.9) | 154 (8.2) |
| Indian | 61 (6.4) | 71 (7.8) | 132 (7.1) |
| Asian/Other | 0 | 1 (0.1) | 1 (0.1) |
| Unknown | 5 (0.5) | 8 (0.9) | 13 (0.7) |
| **Pregnant** | 31 (3.2) | - | 31 (3.2) |
| **Number of comorbidities** |  |  |  |
| No comorbidities | 295 (30.7) | 278 (30.5) | 573 (30.6) |
| 1 comorbidity | 303 (31.6) | 310 (34.0) | 613 (32.7) |
| 2 comorbidities | 234 (24.4) | 196 (21.5) | 430 (23.0) |
| ≥3 comorbidities | 128(13.3) | 129 (14.1) | 257 (13.7) |
| **Comorbidity & risk factors** |  |  |  |
| Hypertension | 330 (34.4) | 339 (37.1) | 669 (35.7) |
| Obesity | 244 (25.4) | 230 (25.2) | 474 (25.3) |
| Diabetes | 204 (21.3) | 214 (23.4) | 418 (22.3) |
| Heart disease | 47 (4.9) | 64 (7.0) | 111 (5.9) |
| High cholesterol | 43 (4.5) | 61 (6.7) | 104 (5.6) |
| HIV | 67 (7.0) | 28 (3.1) | 95 (5.1) |
| Asthma | 58 (6.0) | 34 (3.7) | 92 (4.9) |
| Kidney disease | 11 (1.2) | 19 (2.1) | 30 (1.6) |
| Rheumatological disorder | 18 (1.9) | 9 (1.0) | 27 (1.4) |
| Cancer | 14 (1.5) | 12 (1.3) | 26 (1.4) |
| Chronic lung disease | 11 (1.2) | 14 (1.5) | 25 (1.3) |
| Thyroid disease | 20 (2.1) | 4 (0.4) | 24 (1.3) |
| Depression | 11 (1.2) | 10 (1.1) | 21 (1.1) |
| Other | 126 (13.1) | 97 (10.6) | 223 (11.9) |
| **Health Sector** |  |  |  |
| Private | 687 (71.6) | 724 (79.3) | 1,411 (75.3) |
| Public | 273 (28.4) | 189 (20.7) | 462 (24.7) |
| **Received supplemental oxygen in hospital** | 650 (67.7) | 674 (73.8) | 1,324 (70.7) |
| **Admitted to ICU** | 265 (27.6) | 347 (38.0) | 612 (32.7) |
| **Received invasive mechanical ventilation** | 75 (7.8) | 104 (11.4) | 179 (9.6) |
| **Received at least one dose of a COVID-19 vaccine** | 478 (49.8) | 464 (50.8) | 942 (50.3) |

### Acute and persistent symptoms

Of the 1,873 participants followed up at three months after hospital discharge, 1,672 (89.3%) reported symptoms during the acute phase of their COVID-19 illness. The median number of acute symptoms reported was 4 (IQR 2 – 7). Among the participants reporting acute symptoms, 142 (7.6%) reported one, 211 (11.3%) reported two, 247 (13.2%) reported three and 1,072](57.2%) reported four or more symptoms. The most commonly reported symptoms on admission during the acute COVID-19 illness were fatigue/malaise (56.8%), shortness of breath (50.3%), fever (46.2%), cough (45.3%) and headache (37.2%).

Three months after hospital discharge, 1,249/1,873 (66.7%) participants reported new or persistent symptoms. Among these, 371 (19.8%) reported one, 277 (14.8%) two, 180 (9.6%) three and 421 (22.5%) four or more persistent symptoms. The most commonly reported symptoms three months post-hospital discharge were fatigue (50.3%), shortness of breath (23.4%), confusion or lack of concentration (17.5%), headache (13.8%) and problems seeing/blurred vision (10.1%).

Comparing symptoms at one month and three months after hospital discharge, there was a decline in the prevalence of persistent symptoms from 1,978/2,410 (82.1%) at one month to 1,249/1,873 (66.7%) at three months. However, confusion/lack of concentration, arthralgia, back pain/back ache, and hair loss were more commonly reported at three months compared to at one month. The most commonly reported symptoms at one month were fatigue/malaise (64.9%), shortness of breath (34.8%), headache (20.0%), weakness of the arms or legs (18.8%) and confusion/lack of concentration (16.0%).

### Impact of persistent symptoms on daily living

Most participants reported no problems with mobility, self-care, usual activities, pain/discomfort or anxiety/depression at the three month follow up assessment. However, 219 (11.7%) reported problems with mobility, 255 (13.6%) with performing their usual activities including work, study, housework and family or leisure activities, and 87 (4.6%) with self-care which were all similar for females and males (Table 3).

There were 328 (17.5%) participants who experienced slight, moderate, severe or extreme pain. More females (n=202; 21.0%) reported pain than males (n=225; 13.8%) (p<0.001). There were 222 (11.9%) participants who reported mild anxiety/depression, 112 (6.0%) reported moderate anxiety/depression and 48 (2.6%) reported having severe to extreme anxiety/depression. More females (n=236; 24.6%) than males (n=146; 16.0%) reported having anxiety/depression (p<0.001) (Table 2).

**Table 2.** Prevalence of acute and Post-COVID symptoms reported at one- and three months post-discharge from hospital, PCC Study, South Africa

| **Symptom** | **Acute COVID-19**<br>**n/N (%)** | **One month follow up**<br>**n/N (%)** | **Three months follow up**<br>**n/N (%)** |
| --- | --- | --- | --- |
| Fatigue / Malaise | 1,063 (56.8) | 1,226 (65.5) | 942 (50.3) |
| Shortness of Breath | 942 (50.3) | 864 (46.1) | 439 (23.4) |
| Headache | 696 (37.2) | 406 (21.7) | 258 (13.8) |
| Joint Pain (Arthralgia) | 656 (35) | 145 (7.7) | 175 (9.3) |
| Muscle Aches (Myalgia) | 40 (2.1) | 198 (10.6) | 156 (8.3) |
| Any other symptoms | 170 (9.1) | 148 (7.9) | 149 (8) |
| Chest pain | 475 (25.4) | 280 (14.9) | 132 (7) |
| Dizziness/Light Headedness | 45 (2.4) | 222 (11.9) | 116 (6.2) |
| Cough: Dry Cough | 849 (45.3) | 276 (14.7) | 78 (4.2) |
| Loss of Taste | 111 (5.9) | 162 (8.6) | 51 (2.7) |
| Abdominal pain | 154 (8.2) | 97 (5.2) | 44 (2.3) |
| Loss of Smell | 153 (8.2) | 123 (6.6) | 44 (2.3) |
| Back Pain/Back Ache | 33 (1.8) | 34 (1.8) | 43 (2.3) |
| Cough: With Sputum | 161 (8.6) | 94 (5) | 40 (2.1) |
| Diarrhoea | 293 (15.6) | 73 (3.9) | 38 (2) |
| Skin Rash | 54 (2.9) | 73 (3.9) | 35 (1.9) |
| Fever | 866 (46.2) | 43 (2.3) | 29 (1.5) |
| Nausea / Vomiting | 240 (12.8) | 43 (2.3) | 25 (1.3) |
| Nasal Congestion/Sinusitis | 21 (1.1) | 9 (0.5) | 17 (0.9) |
| Bleeding | 29 (1.5) | 24 (1.3) | 11 (0.6) |
| Loss of Appetite/Anorexia | 429 (22.9) | 12 (0.6) | 10 (0.5) |
| Body Pain/Body Ache | 15 (0.8) | 0 (0) | 2 (0.1) |
| Seizures | 16 (0.9) | 4 (0.2) | 2 (0.1) |

Most participants reported no change in their ability to see, hear, walk, remember and self-care prior to COVID-19 using the UN/Washington score. However, 409 (21.8%), 268 (14.3%) and 179 (9.6%) reported worsening in remembering, walking and seeing, respectively.

Among 1,837 participants followed up at three months, 837 (44.7%) consulted with a general practitioner or primary health care clinic, and 82 (4.4%) were re-admitted to hospital, due to new or persistent symptoms after hospital discharge. There were 36 (1.9%) participants who reported to still be on domiciliary supplemental oxygen three months after hospital discharge and the median time on home oxygen was 21 (IQR 7-30) days.

### Occupation and lifestyle changes

Among all participants, 1,099 (58.7%) were working full-time before contracting COVID-19, while 377 (20.1%), 227 (12.1%) and 69 (3.7%) were retired, unemployed or working part-time, respectively. Among 47 (2.5%) participants that reported a change in occupation at the three months assessment, 15 (31.9%) attributed the changes in occupation to the effects of PCC.

Participants were surveyed about their social habits before and after COVID-19 diagnosis - 81/332 (24.4%) self-reported smokers were smoking less, and 468/712 (65.7%) self-reported alcohol consumers were consuming less alcohol. In addition, 1,065 (56.9%) were eating healthier food, 582 (31.1%) were exercising more, whilst 453 (24.2%) were exercising less.

### Factors associated with Post COVID-19 Condition

On multivariable analysis, factors associated with new or persistent symptoms were age ≥65 years [(aOR 1.62; 95% CI (1.00-2.61)]; female sex [aOR 2.00; 95% CI (1.51-2.65)]; mixed ethnicity [aOR 2.15; 95% CI (1.26-3.66)] compared to black ethnicity; receiving supplemental oxygen during admission [aOR 1.44; 95% CI (1.06-1.97)]; ICU admission [aOR 1.87; 95% CI (1.36-2.57)]; pre-existing obesity [aOR 1.44; 95% CI (1.09-1.91)]; and the presence of ≥4 acute symptoms [aOR 1.94; 95% CI (1.19-3.15)] compared to 0 symptoms during acute COVID-19 phase (Table 4).

**Table 3.**
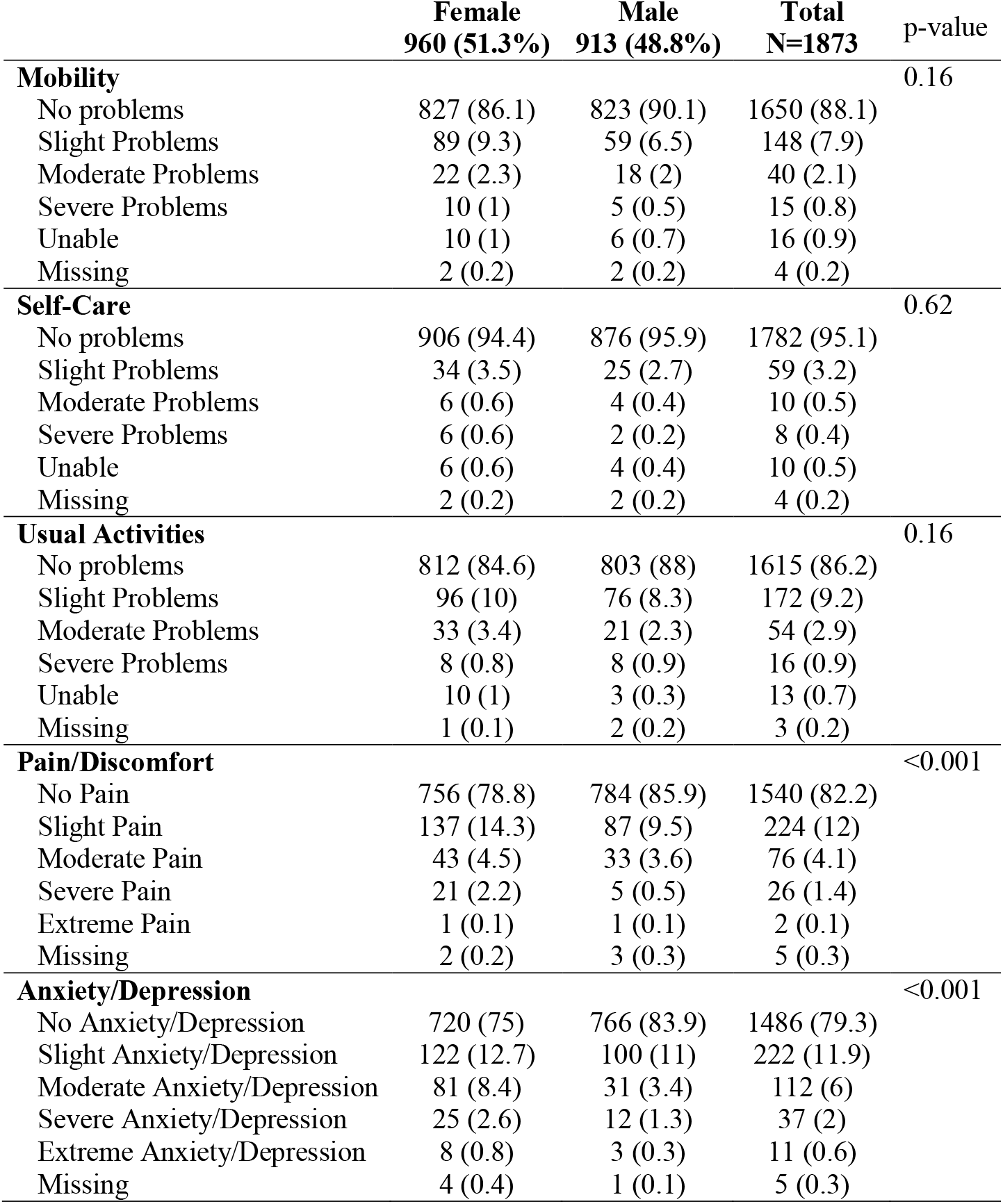
Impact of persistent symptoms on activities of daily living measured using the EuroQol Research Foundation EQ-5D™ tool v2.1, PCC Study, South Africa

|  | **Female**<br>**960 (51.3%)** | **Male**<br>**913 (48.8%)** | **Total**<br>**N=1873** | **p-value** |
| --- | --- | --- | --- | --- |
| **Mobility** |  |  |  | 0.16 |
| No problems | 827 (86.1) | 823 (90.1) | 1650 (88.1) |  |
| Slight Problems | 89 (9.3) | 59 (6.5) | 148 (7.9) |  |
| Moderate Problems | 22 (2.3) | 18 (2) | 40 (2.1) |  |
| Severe Problems | 10 (1) | 5 (0.5) | 15 (0.8) |  |
| Unable | 10 (1) | 6 (0.7) | 16 (0.9) |  |
| Missing | 2 (0.2) | 2 (0.2) | 4 (0.2) |  |
| **Self-Care** |  |  |  | 0.62 |
| No problems | 906 (94.4) | 876 (95.9) | 1782 (95.1) |  |
| Slight Problems | 34 (3.5) | 25 (2.7) | 59 (3.2) |  |
| Moderate Problems | 6 (0.6) | 4 (0.4) | 10 (0.5) |  |
| Severe Problems | 6 (0.6) | 2 (0.2) | 8 (0.4) |  |
| Unable | 6 (0.6) | 4 (0.4) | 10 (0.5) |  |
| Missing | 2 (0.2) | 2 (0.2) | 4 (0.2) |  |
| **Usual Activities** |  |  |  | 0.16 |
| No problems | 812 (84.6) | 803 (88) | 1615 (86.2) |  |
| Slight Problems | 96 (10) | 76 (8.3) | 172 (9.2) |  |
| Moderate Problems | 33 (3.4) | 21 (2.3) | 54 (2.9) |  |
| Severe Problems | 8 (0.8) | 8 (0.9) | 16 (0.9) |  |
| Unable | 10 (1) | 3 (0.3) | 13 (0.7) |  |
| Missing | 1 (0.1) | 2 (0.2) | 3 (0.2) |  |
| **Pain/Discomfort** |  |  |  | <0.001 |
| No Pain | 756 (78.8) | 784 (85.9) | 1540 (82.2) |  |
| Slight Pain | 137 (14.3) | 87 (9.5) | 224 (12) |  |
| Moderate Pain | 43 (4.5) | 33 (3.6) | 76 (4.1) |  |
| Severe Pain | 21 (2.2) | 5 (0.5) | 26 (1.4) |  |
| Extreme Pain | 1 (0.1) | 1 (0.1) | 2 (0.1) |  |
| Missing | 2 (0.2) | 3 (0.3) | 5 (0.3) |  |
| **Anxiety/Depression** |  |  |  | <0.001 |
| No Anxiety/Depression | 720 (75) | 766 (83.9) | 1486 (79.3) |  |
| Slight Anxiety/Depression | 122 (12.7) | 100 (11) | 222 (11.9) |  |
| Moderate Anxiety/Depression | 81 (8.4) | 31 (3.4) | 112 (6) |  |
| Severe Anxiety/Depression | 25 (2.6) | 12 (1.3) | 37 (2) |  |
| Extreme Anxiety/Depression | 8 (0.8) | 3 (0.3) | 11 (0.6) |  |
| Missing | 4 (0.4) | 1 (0.1) | 5 (0.3) |  |

**Table 4.**
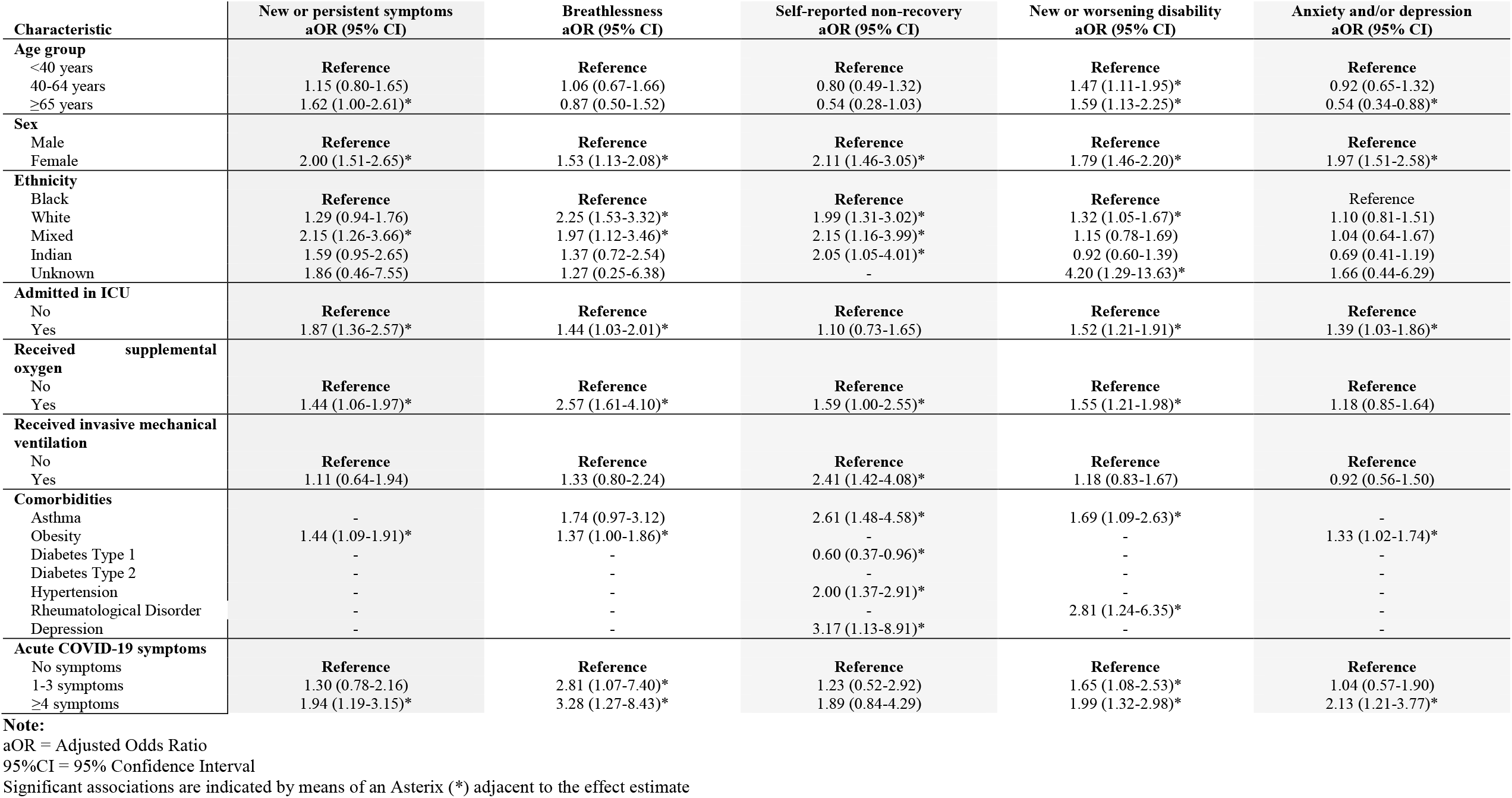
Multivariate analyses of factors associated with five different Post COVID-19 Condition outcomes, PCC Study, South Africa, 1 December 2020 – 23 August 2021, N=1,873.

Factors associated with persistent breathlessness following acute COVID-19 illness were female sex; white and mixed compared to black ethnicity; receiving supplemental oxygen during admission; admission to ICU; pre-existing obesity as well as 1-3 acute symptoms and ≥4 acute symptoms compared to 0 symptoms during the acute COVID-19 phase (Table 4).

Factors associated with self-reported non-recovery were female sex; white, mixed and Indian compared to black ethnicity; receiving supplemental oxygen during admission; receiving mechanical ventilation during admission; pre-existing asthma, hypertension, and anxiety and/or depression (Table 4).

Factors associated with new or worse disability were age between 40 to 64 years and ≥65 years compared to <40 years; female sex; white compared to black ethnicity; receiving supplemental oxygen during admission; ICU admission; pre-existing asthma and rheumatological disorder; and presence of 1-3 acute symptoms and ≥4 acute symptoms compared to 0 symptoms during acute COVID-19 illness (Table 4).

Factors associated with anxiety and/or depression three-months after hospital discharge included female sex; ICU admission; pre-existing obesity; and ≥4 acute symptoms compared to 0 symptoms during acute COVID-19 illness. Being ≥65 years compared to <40 years was protective against anxiety and/or depression (Table 4).

## Discussion

We report new or persistent symptoms were present in two-thirds of participants with confirmed SARS-CoV-2 infection three months post-hospital discharge in this large cohort in South Africa. To our knowledge, this is the first study which describes the prevalence and risk factors for PCC in South Africa and Africa.

We report a similar prevalence of PCC to other studies, that reported prevalence ranging from 50% to 78% (12,13,14,15). Similar to other published studies, the most common PCC symptoms reported by COVID-19 survivors in our cohort were fatigue, shortness of breath, headaches, confusion or lack of concentration, headache, and problems seeing/blurred vision. Studies with shorter follow up (≤ 2 months) reported higher frequency of acute symptoms such as cough, fever and acute gastrointestinal symptoms, which may indicate persistent infection (16,17,18,19), while studies with follow-up beyond three months reported fatigue, shortness of breath and musculoskeletal symptoms more frequently. (12,13,14,15,20,21,22,23)

There are limited studies that reported risk factors for PCC. In this study, we identified an association with age, sex, ethnicity, number of acute COVID-19 symptoms, comorbidities, and acute COVID-19 severity (ICU stay; oxygen; invasive mechanical ventilation) and PCC.

Older adults (over 65 years) had an increased risk for new or persistent symptoms. There is conflicting evidence in the literature on age being a predictor for PCC. Some studies found an association between age younger than 65 years and persistence of symptoms (19,24,25), some reported that increasing age was a risk factor (23,26) while others did not find an association between age and PCC (27).

Female sex was identified as a risk factors for all outcomes, with twice the odds of new or persistent symptoms, self-reported non-recovery, and anxiety and/or depression compared to males. Female sex has been found to have a significant association with PCC in other studies (24,27,28,29,30,31), including associations with fatigue and breathlessness (30), incomplete recovery, and greater disability (24,32). It is important to consider the effect of survival bias in the case of PCC. Males have higher risk of mortality during acute COVID-19 (33) but male sex has not been found to be a significant factor associated with PCC, (27) suggesting that females, typically of younger age, are more likely to survive acute disease, but subsequently experience worse long-term outcomes (32). Furthermore, health-seeking behaviour in males is less than females and males may be less inclined to report persistence of poor health (32). However, other factors such as impact of sex hormones on the risk of developing PCC needs further exploration.

Our study showed a significant association between non-black ethnicity and PCC outcomes in South Africa. A meta-analysis (31) including ten studies based predominantly in Europe also found that black and South Asian ethnicity were protective against PCC. There is limited data available on associations of ethnicity with PCC, and the reasons for differences in persistent symptoms by ethnicity is unclear and will require further investigation.

Admission to ICU was significantly associated with all outcomes except for self-reported non-recovery. Post-intensive care syndrome (PICS), which is a collection of physical, mental and emotional post-ICU persistent sequelae (34,35,36,37,38). The PCC persistent symptoms identified amongst the post-ICU survivors may overlap with or be further compounded by PICS. Participants who received supplemental oxygen during hospital admission also showed significant associations with all outcomes except for anxiety and/or depression; however, mechanical ventilation during admission only showed significant association with self-reported non-recovery. This is in keeping with the outcomes in a study by Scott *et al*. (24) who found that participants who had received invasive ventilation were nearly four times more likely to report incomplete recovery compared to those who had not required any supplemental ventilation or oxygen.

Pre-existing comorbidities such as asthma, diabetes, hypertension, rheumatological disorders, depression/anxiety and obesity were associated with PCC. Previous studies have also found an increased risk of PCC in people with certain pre-existing comorbidities (26,31,32), such as asthma (27,31), hypertension (26), obesity (26,31), neuro-psychological (26,31), and immunosuppressive conditions (26). The pathophysiology of PCC is not well understood, with multiple hypotheses attempting to describe the potential mechanisms based on previous coronavirus outbreaks and other RNA viruses. Some have suggested that PCC may be the result of tissue/organ damage (39,40), or inflammatory and immune pathways dysfunction (including chronic inflammation, hyperactive immune cells and autoimmunity as a result of molecular mimicry) (39,40). In addition, it is theorised that PCC could be due to viral persistence (39), reactivation of latent pathogens (i.e. herpesviruses, Epstein Barr virus) (39) and disruption of commensal microbiomes/virome communities (40). Other plausible mechanisms included clotting/coagulation issues and dysfunctional brainstem/vagus nerve signalling (40). It is plausible that patients with underlying comorbidities admitted with COVID-19 would be at greater risk of PCC because of pre-existent tissue damage, and more severe COVID-19 illness associated with longer hospital stays, more complications and the likelihood of receiving oxygen, invasive ventilation, steroids and other treatment.

Participants with a greater number of symptoms during acute COVID-19, were more likely to report persistent symptoms, breathlessness, disability, and anxiety/depression. This is in line with a study by Sudre *et al* (27) that found that participants who presented with more than five symptoms in the first week of acute illness had a significantly increased risk of PCC.

We report a decline in the prevalence of PCC between the one and three month assessment (82% to 67%), consistent with other studies (3). A longitudinal study by *Huang, L et al*. (41) of previously hospitalised COVID-19 patients found that the prevalence of new or persistent symptoms declined from 68% at six months to 49% at 12 months, suggesting that there is a steady resolution of symptoms with time. However, a prolonged recovery time may still have significant impact on the individual and families, as well as wider socioeconomic and public health implications.

### Strengths and limitations

This is a large nationally representative longitudinal cohort study and the first of its kind in South Africa and Africa. As part of the global ISARIC collaboration, we used standardised and validated tools, which allow comparison across participating countries. The alignment to the DATCOV hospital surveillance system and national SARS-CoV-2 case list allowed us to identify potential participants, and to link key demographic and clinical data related to their hospital admission. These are preliminary results of an ongoing study. The study will continue to follow up participants until 12 months after hospital discharge.

The study had several limitations. Firstly, the study was limited to participants who were hospitalised with SARS-CoV-2. In the future it would be of interest to include those who were not hospitalised and participants with other respiratory infections who tested negative for SARS-CoV-2. The inclusion of only previously hospitalised participants may result in over-reporting of symptoms not specifically attributable to acute COVID-19 or PCC. Some of the symptoms may relate to PICS or complications arising directly from hospitalisation. Secondly, all participants were enrolled through a telephone assessment, limiting the enrolment to those that had a phone number recorded. This may explain the greater proportion of individuals from higher socio-economic strata that were enrolled. Thirdly, participants who experienced symptoms may have been more likely to participate than those who did not. The possibility of recall and response bias and the subjective rating of symptoms may affect the reporting. However, our comparison of demographic characteristics of the participants and the hospitalised participants from which the sample was drawn, found similar distribution by sex, ethnicity, and province, except for a higher proportion of study participants from the private sector and aged 40-64 years (related to availability of contact details and willingness to participate), and with more severe acute illness (related to underreporting in the DATCOV system) (Supplementary Table 1).

## Conclusions

The findings of this national study demonstrates a high prevalence of persistent symptoms at three months post-discharge from hospital following SARS-CoV-2 infection, with impact on daily living, functioning and occupation. The high burden of PCC is concerning for South Africa, from an individual and public health perspective, due to the potential additional burden on an already overwhelmed health care system, reduced work productivity, and increased need for economic support. These findings will inform public health measures, including identifying those at increased risk of developing PCC, and providing patient support and information. Moreover, the findings will inform development of clinical pathways and guidelines for the care of these patients, and health service planning. Long-term follow-up of this cohort will provide further insights into the evolution of PCC in South Africa. Future analyses will also allow us to assess the effect of COVID-19 vaccination on persistent symptoms, and to compare PCC between South Africa’s first, second and third wave, when different virus lineages predominated.

## Data Availability

All data produced in the present study are available upon reasonable request to the authors.

## Acknowledgements

We would like to acknowledge all the members of the research team (namely Ashrina Kandier, Bawinile Hlela, Bibianna Chikowore, Claudette Kibasomba, Dorcas Magorimbo-Njanjeni, Jeniffer Nagudi, Kadija Shangase, Khutso Maphoto, Lilford Lesabe, Lindelwa Ngobeni, Manana Sibanda, Menzi Mbonambi, Molebogeng Moyo, Ncamsile Mavundla, Okaeng Plaatjie, Salaminah Mhlanga, Sinalo Gqunu, Tasmeeya Moola, Thandeka Kosana and Zelna Jacobs) responsible for the recruitment of participants into the study and subsequent follow up surveys. We are grateful to the Bill and Melinda Gates Foundation (especially Georgina Murphy and Keith Klugman) for their continued support of the project. We thank the ISARIC team (especially Daniel Plotkin, Laura Merson and Louise Sigfrid) for their help in developing the CRF and supporting the REDCap database. Moreover, members of the Long COVID support group who informed the CRF. Finally, we would like to acknowledge all members of the NICD and DATCOV Team (especially Richard Welch) who have provided technical support and assisted with data management and express gratitude to all the patients who participated in the contribution to the understanding of Post COVID-19 Condition.

## Supplemental Material

### Supplementary Methods: Statistical Analysis

The outcome variables were obtained as follows:

- New or persistent symptoms: obtained from the number of persistent symptoms the participants have. Those who did not have any symptoms were classified as “No persistent symptoms” and those with 1 or more symptoms were classified as “New or persistent symptoms”.
- Self-reported non-recovery: obtained from the Likert scale question; “Do you feel fully recovered from your COVID-19?”. The five possible responses to this question were collapsed into two categories “Recovered” and “Not recovered”. The response “Neither agree or disagree” was treated as neutral and excluded from the analysis.
- New or worsening breathlessness: obtained from the question “How breathless you feel TODAY and how breathless you felt BEFORE your COVID-19 illness”. Responses were coded from a value of 1 to 5 representing less severe to very severe breathlessness. The output obtained from difference between breathlessness before and breathlessness after (breathless before – breathless after) was classified into two categories. A difference greater or equal to zero was classified as “No new or worsening breathlessness” = 0 (breathing better or no change in breathlessness) and a difference of less than zero was classified as “New or worsening breathlessness” = 1.
- New/worse disability: constructed from the Washington short set tool (11) which includes questions on changes in vision, hearing, mobility, remembering, self-care and communication (comparing before and after contracting COVID-19). Responses to these questions were coded from 1 to 4 representing no difficulty to inability to function. Change in any disability was classified as “Better”, “No Change” and “Worse/New disability”. The outcome variable “New/worse disability” was constructed if worse change or a new disability was noted in any one of the stated Washington disability categories (“No disability” = 0 and “At least one disability” = 1).
- Anxiety/Depression: constructed from the EuroQol Research Foundation EQ-5D™ tool v2.1 (9). The responses to the Likert scale Anxiety/Depression question were collapsed into two categories, “anxiety/ depression” if there was any level of anxiety or depression reported and “no anxiety/ depression”.

**Supplementary Table 1.**
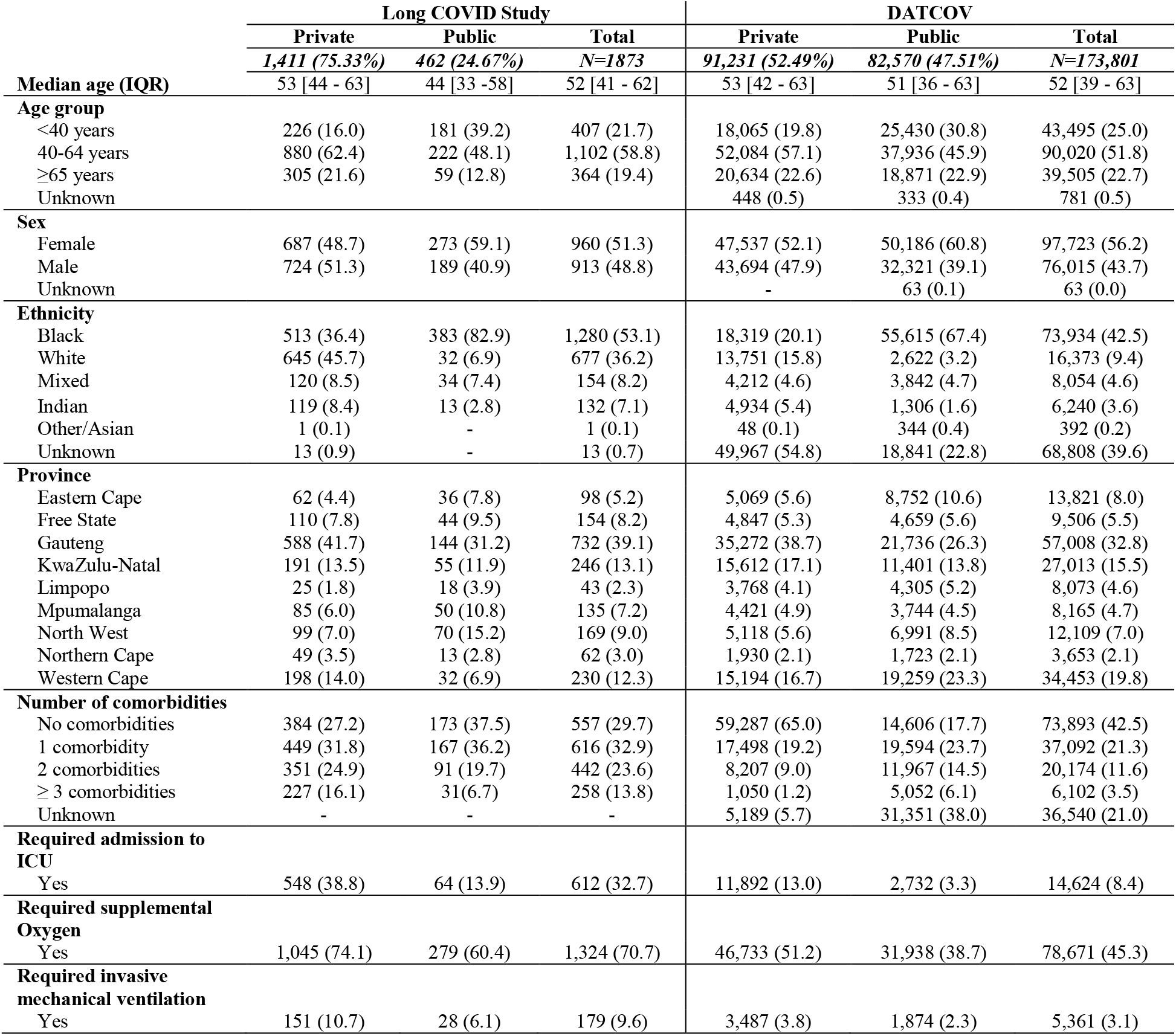
Characteristics of study patients and hospitalised patients in DATCOV, a hospital-based COVID-19 surveillance network coordinated by the National Institute for Communicable Diseases, South Africa.

|  | Long COVID Study |  |  | DATCOV |  |  |
| --- | --- | --- | --- | --- | --- | --- |
|  | Private | Public | Total | Private | Public | Total |
|  | **1,411 (75.33%)** | **462 (24.67%)** | **N=1873** | **91,231 (52.49%)** | **82,570 (47.51%)** | **N=173,801** |
| **Median age (IQR)** | 53 [44 - 63] | 44 [33 -58] | 52 [41 - 62] | 53 [42 - 63] | 51 [36 - 63] | 52 [39 - 63] |
| **Age group** |  |  |  |  |  |  |
| <40 years | 226 (16.0) | 181 (39.2) | 407 (21.7) | 18,065 (19.8) | 25,430 (30.8) | 43,495 (25.0) |
| 40-64 years | 880 (62.4) | 222 (48.1) | 1,102 (58.8) | 52,084 (57.1) | 37,936 (45.9) | 90,020 (51.8) |
| ≥65 years | 305 (21.6) | 59 (12.8) | 364 (19.4) | 20,634 (22.6) | 18,871 (22.9) | 39,505 (22.7) |
| Unknown |  |  |  | 448 (0.5) | 333 (0.4) | 781 (0.5) |
| **Sex** |  |  |  |  |  |  |
| Female | 687 (48.7) | 273 (59.1) | 960 (51.3) | 47,537 (52.1) | 50,186 (60.8) | 97,723 (56.2) |
| Male | 724 (51.3) | 189 (40.9) | 913 (48.8) | 43,694 (47.9) | 32,321 (39.1) | 76,015 (43.7) |
| Unknown |  |  |  | - | 63 (0.1) | 63 (0.0) |
| **Ethnicity** |  |  |  |  |  |  |
| Black | 513 (36.4) | 383 (82.9) | 1,280 (53.1) | 18,319 (20.1) | 55,615 (67.4) | 73,934 (42.5) |
| White | 645 (45.7) | 32 (6.9) | 677 (36.2) | 13,751 (15.8) | 2,622 (3.2) | 16,373 (9.4) |
| Mixed | 120 (8.5) | 34 (7.4) | 154 (8.2) | 4,212 (4.6) | 3,842 (4.7) | 8,054 (4.6) |
| Indian | 119 (8.4) | 13 (2.8) | 132 (7.1) | 4,934 (5.4) | 1,306 (1.6) | 6,240 (3.6) |
| Other/Asian | 1 (0.1) | - | 1 (0.1) | 48 (0.1) | 344 (0.4) | 392 (0.2) |
| Unknown | 13 (0.9) | - | 13 (0.7) | 49,967 (54.8) | 18,841 (22.8) | 68,808 (39.6) |
| **Province** |  |  |  |  |  |  |
| Eastern Cape | 62 (4.4) | 36 (7.8) | 98 (5.2) | 5,069 (5.6) | 8,752 (10.6) | 13,821 (8.0) |
| Free State | 110 (7.8) | 44 (9.5) | 154 (8.2) | 4,847 (5.3) | 4,659 (5.6) | 9,506 (5.5) |
| Gauteng | 588 (41.7) | 144 (31.2) | 732 (39.1) | 35,272 (38.7) | 21,736 (26.3) | 57,008 (32.8) |
| KwaZulu-Natal | 191 (13.5) | 55 (11.9) | 246 (13.1) | 15,612 (17.1) | 11,401 (13.8) | 27,013 (15.5) |
| Limpopo | 25 (1.8) | 18 (3.9) | 43 (2.3) | 3,768 (4.1) | 4,305 (5.2) | 8,073 (4.6) |
| Mpumalanga | 85 (6.0) | 50 (10.8) | 135 (7.2) | 4,421 (4.9) | 3,744 (4.5) | 8,165 (4.7) |
| North West | 99 (7.0) | 70 (15.2) | 169 (9.0) | 5,118 (5.6) | 6,991 (8.5) | 12,109 (7.0) |
| Northern Cape | 49 (3.5) | 13 (2.8) | 62 (3.0) | 1,930 (2.1) | 1,723 (2.1) | 3,653 (2.1) |
| Western Cape | 198 (14.0) | 32 (6.9) | 230 (12.3) | 15,194 (16.7) | 19,259 (23.3) | 34,453 (19.8) |
| **Number of comorbidities** |  |  |  |  |  |  |
| No comorbidities | 384 (27.2) | 173 (37.5) | 557 (29.7) | 59,287 (65.0) | 14,606 (17.7) | 73,893 (42.5) |
| 1 comorbidity | 449 (31.8) | 167 (36.2) | 616 (32.9) | 17,498 (19.2) | 19,594 (23.7) | 37,092 (21.3) |
| 2 comorbidities | 351 (24.9) | 91 (19.7) | 442 (23.6) | 8,207 (9.0) | 11,967 (14.5) | 20,174 (11.6) |
| ≥ 3 comorbidities | 227 (16.1) | 31(6.7) | 258 (13.8) | 1,050 (1.2) | 5,052 (6.1) | 6,102 (3.5) |
| Unknown | - | - | - | 5,189 (5.7) | 31,351 (38.0) | 36,540 (21.0) |
| **Required admission to ICU** |  |  |  |  |  |  |
| Yes | 548 (38.8) | 64 (13.9) | 612 (32.7) | 11,892 (13.0) | 2,732 (3.3) | 14,624 (8.4) |
| **Required supplemental Oxygen** |  |  |  |  |  |  |
| Yes | 1,045 (74.1) | 279 (60.4) | 1,324 (70.7) | 46,733 (51.2) | 31,938 (38.7) | 78,671 (45.3) |
| **Required invasive mechanical ventilation** |  |  |  |  |  |  |
| Yes | 151 (10.7) | 28 (6.1) | 179 (9.6) | 3,487 (3.8) | 1,874 (2.3) | 5,361 (3.1) |

